# Toward optimal moxifloxacin dosing in tuberculous meningitis: a translational physiologically based pharmacokinetic modeling approach

**DOI:** 10.1101/2025.11.27.25341172

**Authors:** Ming Sun, Katie Lynch, Theis Mariager, Jacob Bodilsen, Roland Nau, Rob C. van Wijk, Martijn L. Manson, Elizabeth C.M. de Lange, Tingjie Guo

## Abstract

**Background:** Tuberculous meningitis (TBM) is a severe central nervous system (CNS) infection with high mortality. Moxifloxacin shows potent anti-mycobacterial activity and favorable CNS penetration. However, optimal dosing remains uncertain, particularly with rifampicin, which markedly reduces moxifloxacin exposure. This study aimed to determine optimal moxifloxacin dosing for TBM by accounting for regional CNS pharmacokinetics (PK) and rifampicin co-administration using a cross-species translational physiologically based pharmacokinetic (PBPK) model.

**Methods:** A PBPK model was developed using high-resolution plasma and CNS microdialysis data from pigs to capture CNS physiology and moxifloxacin distribution. The model was translated to humans using literature-derived physiological parameters and allometric scaling and validated with plasma and CNS PK data from healthy subjects and TBM patients. Simulations of once-daily moxifloxacin doses (400 to 1000 mg), with and without rifampicin, were evaluated using the unbound fAUC0-24/MIC target of 53.

**Results:** The model accurately reproduced observed moxifloxacin concentrations in porcine and human CNS compartments. Simulations showed regional PK differences, with highest concentrations in the subarachnoid space and cisterna magna and lowest in brain extracellular fluid. Rifampicin reduced mean CNS exposure by 26%. Without rifampicin, target attainment was achieved at 800 mg once daily, while no simulated regimen reached target levels with rifampicin co-administration.

**Conclusion:** These results suggest that an 800 mg once-daily moxifloxacin regimen may provide adequate CNS exposure in TBM patients not receiving rifampicin. For patients treated with rifampicin, a higher dose might be necessary to achieve therapeutic targets.

## 1 Introduction

Tuberculous meningitis (TBM) is the most severe manifestation of central nervous system (CNS) tuberculosis (TB), associated with high mortality and long-term neurological disability despite treatment (1, 2). The World Health Organization (WHO) recommends an initial four-drug regimen consisting of isoniazid, rifampicin, pyrazinamide, and ethambutol, but its efficacy is increasingly undermined by multidrug resistance and treatment-related adverse effects (3, 4). Recent efforts on investigating intensified rifampicin dosing or adjunctive linezolid have shown limited success in improving TBM outcomes (5, 6). Moxifloxacin, a fourth-generation fluoroquinolone, has potent bactericidal activity against *Mycobacterium tuberculosis* (Mtb) (7, 8) and favorable CNS penetration (9), making it a promising adjunct or alternative in TBM regimen, particularly for drug-resistant or refractory cases (10). It was also essential for the pulmonary tuberculosis treatment shortening from 6 to 3 months together with rifapentine (11). Since rifampicin is the cornerstone drug that commonly used in the combination therapy, it can interact with other drugs such as moxifloxacin by inducing phase II drug-metabolizing enzymes to reduce their systemic concentrations (12). This drug-drug interaction complicates optimization of combination regimens and underscores the need for a deeper understanding of moxifloxacin pharmacokinetics (PK) in the CNS during rifampicin co-administration.

Available human PK data are limited to sparse cerebrospinal fluid (CSF)-to-plasma ratios or isolated case reports, which do not reliably reflect drug concentrations at key CNS sites involved in TBM pathogenesis (4, 13–15). Mtb is believed to first cross the blood-brain barrier (BBB), reside the parenchyma (forming Rich foci), and later enter into CSF compartments, particularly the subarachnoid space (SAS), eventually causing inflammation (16, 17). Thus, understanding regional CNS PK exposure beyond CSF alone is critical to improve the current treatment.

Animal models remain essential for studying CNS PK to overcome the limitations of direct human sampling. However, translating findings from animal to human requires a mechanistic approach that accounts for interspecies physiological differences. Physiologically based pharmacokinetic (PBPK) modeling provides a robust framework for achieving this goal. Such models incorporate species-specific anatomy and physiology to simulate regional drug distribution, with human parameters obtained from literature or derived via allometric scaling (18) (19). By further integrating TBM pathology, CNS-focused PBPK models are capable of mechanistically predicting drug exposure across relevant CNS compartment in TBM patients, including brain extracellular fluid and multiple CSF compartments(16, 17). To date, no study has applied such a framework to moxifloxacin CNS distribution in TBM patients.

In this study, we aimed to investigate optimal moxifloxacin treatment for TBM by mechanistically characterizing its regional CNS distribution with and without rifampicin using a translational PBPK modeling approach informed by detailed porcine data.

## 2 Method

### 2.1 Data Sources

#### 2.1.1 Porcine PK Data

Preclinical data from a porcine model (20) served as the foundation for developing the CNS PBPK model. The study included six female pigs (Danish Landrace breed, 3 months old, weighing 38–48 kg) receiving a single moxifloxacin intravenous (IV) dose of 6 mg/kg. Unbound moxifloxacin concentrations were obtained from plasma and multiple CNS regions including brain extracellular fluid (brainECF), brain ventricles (VENs), cisterna magna (CM), and lumbar site using microdialysis. Extensive samples were collected at 40, 80, 120, 160, 200, 240, 300, 360, 420, and 480 minutes post-dose.

#### 2.1.2 Human PK Data - Non-inflamed Meninges

PK data from a published study involving patients with non-inflamed meninges undergoing urological surgery (21) were extracted and used to evaluate the translational performance from pigs to humans of the developed PBPK model. In this study, 50 patients received a single oral dose of 400 mg moxifloxacin prior to surgery. Moxifloxacin concentrations were measured in both plasma and lumbar CSF, with patients divided into five groups (n=10) according to predefined sampling intervals: group I (0.5–1 h), group II (1–2 h), group III (2–4 h), group IV (4–6 h), and group V (6–8 h). These data provided reference concentrations for model validation in the absence of CNS inflammation.

#### 2.1.3 Human PK Data - TBM Patients

PK data from six studies involving patients with tuberculous meningitis (TBM) were extracted and used to validate the predictive performance of the PBPK model under TBM conditions in humans (Table S1). These studies provided clinical data on moxifloxacin concentrations in plasma and CSF of TBM patients, which was used to evaluate the model’s applicability in the disease condition (4, 14, 15, 22–24).

### 2.2 CNS PBPK model development and translation

#### 2.2.1 Porcine CNS PBPK Model

Plasma PK was modeled using an empirical compartmental model structure. One-, two-, and three-compartment plasma models were evaluated, and visual inspection of the fitted concentration–time profiles indicated that the two-compartment model provided the best agreement with the observed plasma data. A CNS PBPK model was then constructed to describe moxifloxacin distribution in pigs across CNS compartments. The CNS PBPK model structure included brain ECF, brain ventricles, cisterna magna, and SAS, with drug exchange governed by permeability and CSF flow parameters (Figure 1). Pig-specific CNS physiological parameters were sourced from literature where possible (Table S2), with any missing values estimated through model fitting to the porcine PK dataset (Table S3).

**Figure 1:**
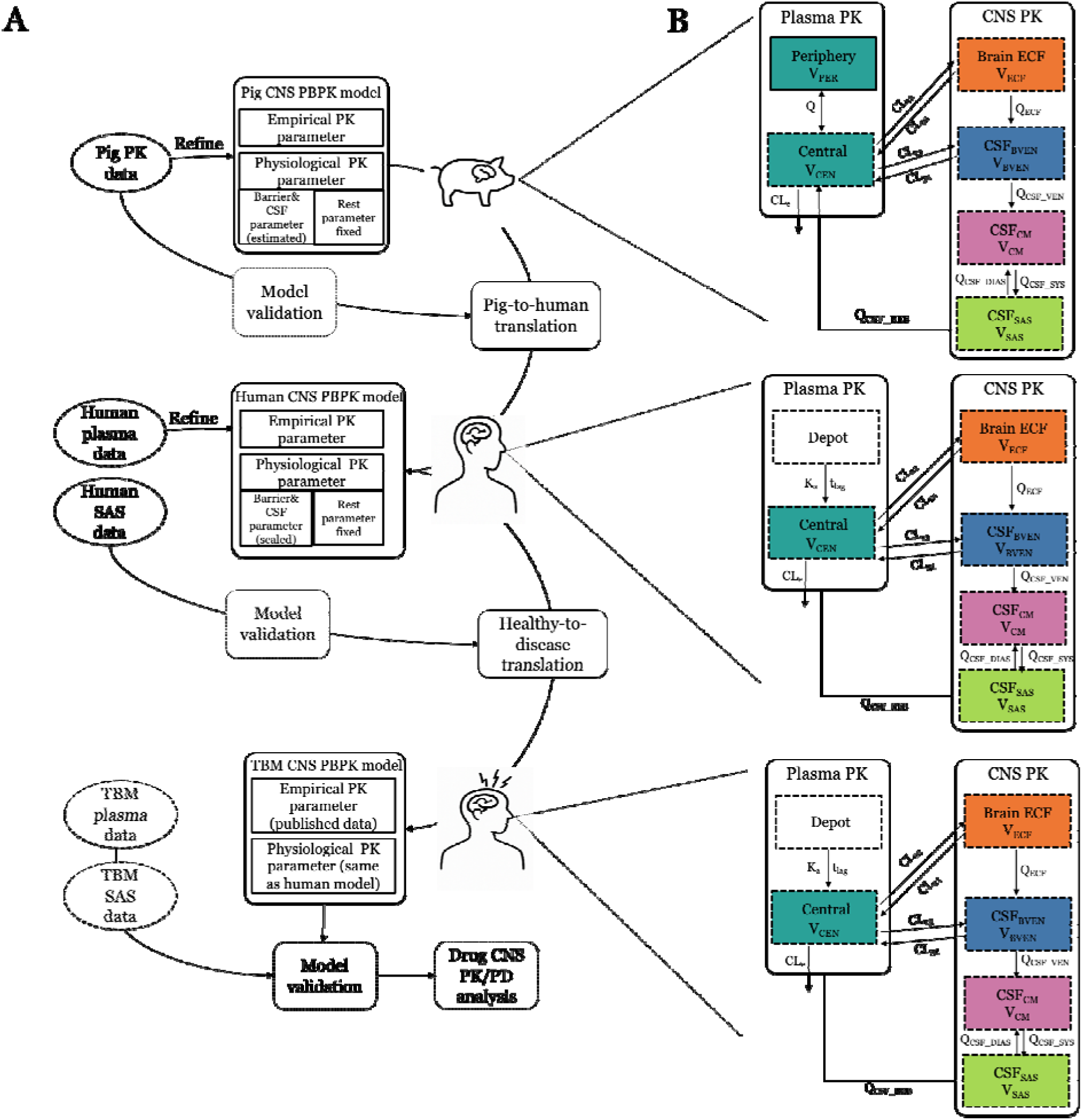
Translational PBPK modeling workflow and CNS model structure for moxifloxacin. (A) Model development and translation of the moxifloxacin PBPK model. A healthy-pig PBPK model was built with systemic circulation PK parameters (the absorption rate constant (K_a_), central compartment volume (V_CEN_), peripheral volume (V_PER_), systemic clearance (CL_e_), inter-compartmental clearance (Q), estimated CNS barrier & CNS flow related parameters (CL_12_, CL_21_, CL_13_, CL_31_, QECF, QCSF_VEN_, QCSF_DIAS_, QCSF_SYS_, and QCSF_REB_), and fixed physiological volumes (VECF, VBVEN, VCM, VSAS) of CNS compartments. For human translation, pig physiological parameters were replaced by literature-reported human value where available, while unmeasured parameters (including BBB/BCSFB barrier and CSF related parameters) were allometrically scaled; empirical PK parameters were then re-estimated from plasma profiles of non-meningitis subjects, and the resulting model were validated against subarachnoid-space (SAS) concentrations. The healthy-human model was then adapted to tuberculous meningitis (TBM) b replacing plasma model parameters under healthy condition with diseased values and was validated against drug plasma and SAS concentrations of TBM patients, yielding a disease-specific PBPK model. (B) Schematic of the CNS PBPK submodel structures. A central (plasma) compartment (V_CEN_) is linked either to a peripheral tissue compartment (V_PER_, Q) or to an absorption depot (k_a_, t_lag_), with elimination CL_e_. Drug distributes between plasma and brain extracellular fluid (ECF; V_ECF_) via barrier clearances (CL_12_, CL_21_, CL_13_, CL_31_). From ECF, moxifloxacin can cross three serial CSF compartments i.e. brain ventricles (V_BVEN_), cisterna magna (V_CM_) and SAS (V_SAS_) driven by unidirectional flows (QECF, QCSF_VEN_) and bidirectional CSF flows (QCSF_DIAS_, QCSF_SYS_) with recirculation (QCSF_REB_) back to plasma. Arrows indicate the direction of drug transport.

#### 2.2.2 Human CNS PBPK Model

In order to translate the porcine model to human, species-specific physiological parameters (e.g., brain volume, surface area, CSF volume) of the developed porcine PBPK model were replaced with human values obtained from literatures. BBB/BCSFB-related parameters (CL_12_, CL_21_, CL_13_, CL_31_), and CNS flow parameters (QECF, QCSF_VEN_, QCSF_DIAS_, QCSF_SYS_, QCSF_REB_), were scaled using allometric methods (Equation 1 and 2), due to the absence of direct human literature values (Figure 1):

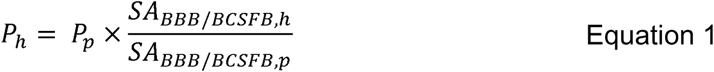

Where SA_BBB/BCSFB_ stands for the surface area of the BBB or blood-CSF barrier (BCSFB). Typical values of SA_BBB/BCSFB_ for pigs and humans (Table S4) were used to calculate surface area ratios. BBB/BCSFB-crossing related parameters were scaled using Equation 1 based on the surface area ratio, given the conserved expression levels of most active transporters in the plasma membrane fractions of the BBB and BCSFB between pig and human (25–27).

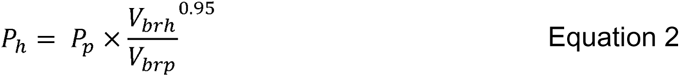

Where Vbrh/brp stands for the brain volume of human or pig. CNS flow-related parameters were scaled from pig to human using Equation 2 based on human-to-pig brain volume (V_br_) ratio using an average exponent of 0.95 from a previous cross-species cortical blood flow scaling practices (28). Using an exponent greater than the conventional 0.75 reflects the brain’s disproportionately high metabolic rate compared to whole-body processes (29). Physiological parameters such as brain weight, volume, and surface area were fixed using published human physiological values (Table S4).

Plasma PK parameters, including lag time (t_lag_), K_a_, V_CEN_, and CL_e_ (Table S3), were determined by fitting published mean plasma concentration data from human subjects without meningitis (21) using a one-compartment model. The plasma unbound fraction (f_u_) was fixed as a reported value of 50% (30, 31).

#### 2.2.3 Model Adaptation for TBM Conditions

CNS related parameters, including CL_12_, CL_21_, CL_13_, CL_31_, were assumed to be identical between TBM patients and non-TBM patients, given moxifloxacin’s favorable CNS penetration under non-inflamed conditions and the likely minimal increase in CNS clearance associated with inflammation (21). Moreover, there were no quantitative data available to justify adjusting the other CNS related parameters (QECF, QCSF_VEN_, QCSF_DIAS_, QCSF_SYS_, and QCSF_REB_). To account for the effect of rifampicin on moxifloxacin plasma clearance (32) as well as the effect of systemic infection, we collected plasma PK parameters for both moxifloxacin alone and in combination with rifampicin (Table S3) from the tuberculosis patients (33).

### 2.3 CNS PBPK Model Diagnostics and Validation

#### 2.3.1 Pig model diagnostics

Model performance in pigs was assessed by comparing predicted against observed concentration-time profiles in all compartments. Goodness-of-fit plots and visual predictive check (VPC) (34) were used to evaluate prediction accuracy at both population and individual levels. Residual scatter plots were used to assess model bias.

#### 2.3.2 Validation in Humans with Non-inflamed Meninges

Model predictions of moxifloxacin concentrations in the human SAS were evaluated using root mean square error (RMSE) to quantify accuracy against observed data from non-meningitis patients. To assess the robustness of our translational approach, we additionally compared our predictions to those generated using alternative allometric scaling methods for barrier permeability and CSF flow parameters (Table S4).

#### 2.3.3 Validation in TBM Patients

The adapted TBM model was validated by comparing predicted PK metrics including the area under the concentration–time curve over 24□hours (AUC_0-24_), maximum concentration (C_max_), half-life (t_1/2_) and full concentration-time profiles, against observed data from published TBM studies (Table 1). These comparisons assessed the model’s ability to capture moxifloxacin behavior in the presence of disease-related drug CNS PK alteration.

### 2.4 Simulation Studies

To assess regional CNS PK in comparison to plasma, moxifloxacin steady-state concentrations were simulated for a standard oral dosing regimen of 400□mg once daily across multiple CNS compartments: plasma, brain ECF, brain VENs, CM, and SAS.

To assess moxifloxacin efficacy against Mtb, the ratio of the AUC_0-24_ to the minimal inhibitory concentration (MIC) was applied as a predictive index, with thresholds of at least 100 based on total drug concentrations (35) and 53 for unbound concentrations (36). The unbound AUC_0-24_ was simulated at steady state following oral doses of 400 mg, 600 mg, 800 mg, and 1000 mg once daily across all the compartments. The clinical breakpoint MIC for Mtb was set at 0.5 mg/L (37). The resulting unbound AUC_0-24_/MIC ratios for each compartment were then compared against the efficacy threshold of 53.

All simulations were conducted considering two scenarios, with and without rifampicin co-administration.

### 2.5 Software and Tools

The observed concentration data were extracted using WebPlotDigitizer version 4.2. Parameter estimations were performed in Monolix 2024R1(38). Data simulations were conducted by the R package rxode2 version 4.1.0 (39). All visualizations, including concentration-time profiles, goodness-of-fit plots, and residual scatter plots, were generated using the R package ggplot2 version 3.5.1.

## 3 Results

### 3.1 Model Performance in Pigs

The porcine CNS PBPK model accurately captured moxifloxacin concentration-time profiles across plasma and CNS compartments. Predicted median concentrations and 90% prediction intervals aligned closely with observed data from six individual pigs (Figure 2). Goodness-of-fit plots at both individual and population levels showed overall agreement between observed and predicted values, however, observed concentrations from pig ID3 in the brainVENs, CM, and SAS compartments were higher than predicted at population levels (Figure S1). Individual fits confirmed accurate tracking of observed concentrations across compartments (Figure S2). Visual predictive checks (Figure S3) indicated good model performance, although residual diagnostics (Figure S4) revealed a time-dependent pattern in plasma, brainECF, and CM, suggesting mild residual error drift over the sampling interval. Overall, these findings support the reliability of the porcine model in characterizing moxifloxacin CNS distribution.

**Figure 2:**
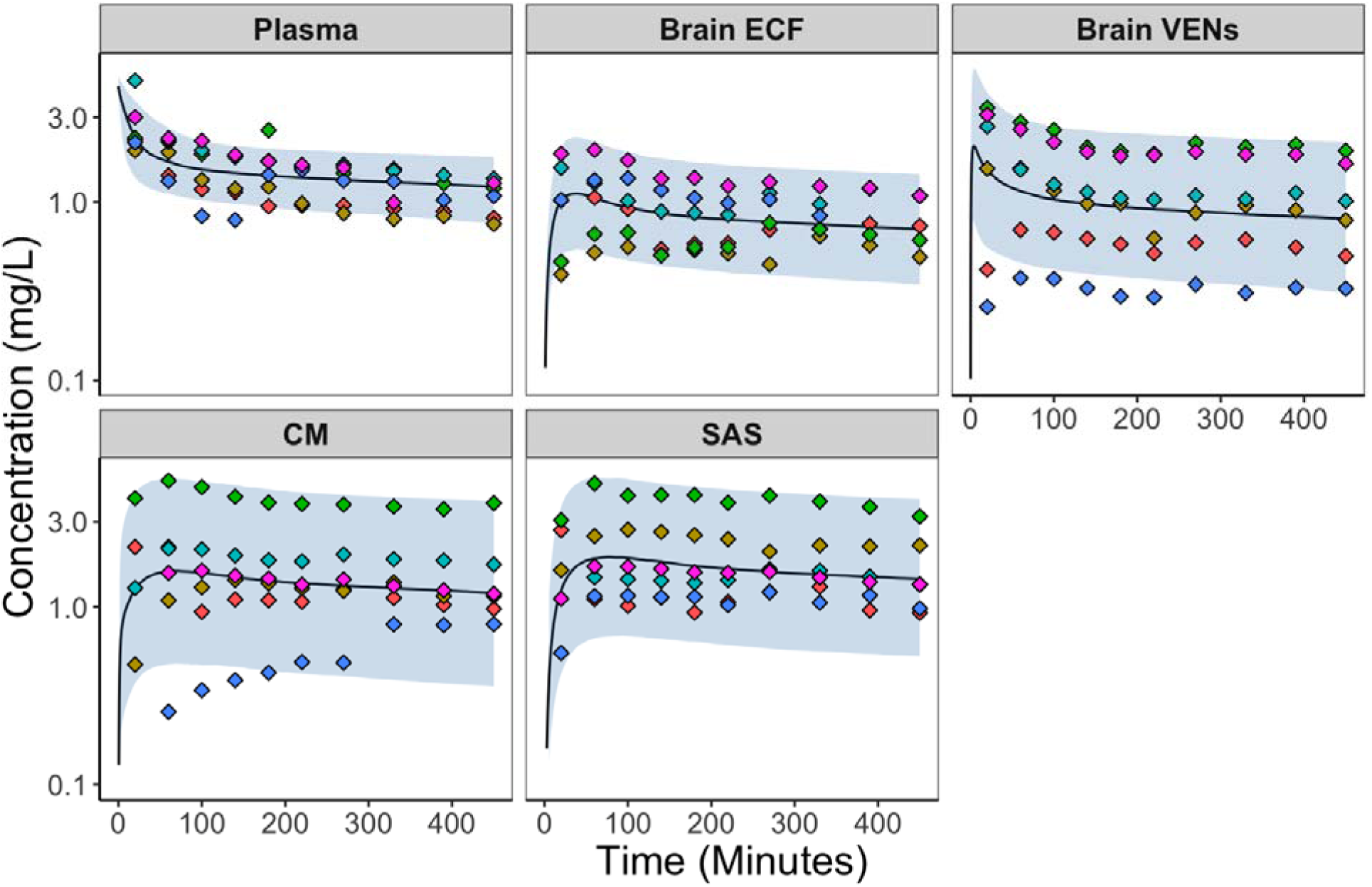
Model-predicted moxifloxacin concentration-time profiles in plasma, brain extracellular fluid (ECF), brain VENs (ventricles), cisterna magna (CM), and subarachnoid space (SAS) in pigs. Observed data from six pigs (20) (colored points) and 90% prediction intervals (shaded regions) were shown on log Y scale.

### 3.2 Model Validation in Humans with Non-inflamed Meninges

The translated human CNS model showed good predictive accuracy in non-meningitis patients receiving moxifloxacin monotherapy. Predicted moxifloxacin concentrations in the SAS were consistent with observed data across sampling intervals (Figure 3). When compared to other published scaling methods (Table S4), our approach yielded the closest match to clinical observations, whereas alternative scaling methods produced larger deviations (RMSE 0.97 vs. RMSE 1.2-1.5; Figure S5), supporting the suitability of our scaling strategy.

**Figure 3:**
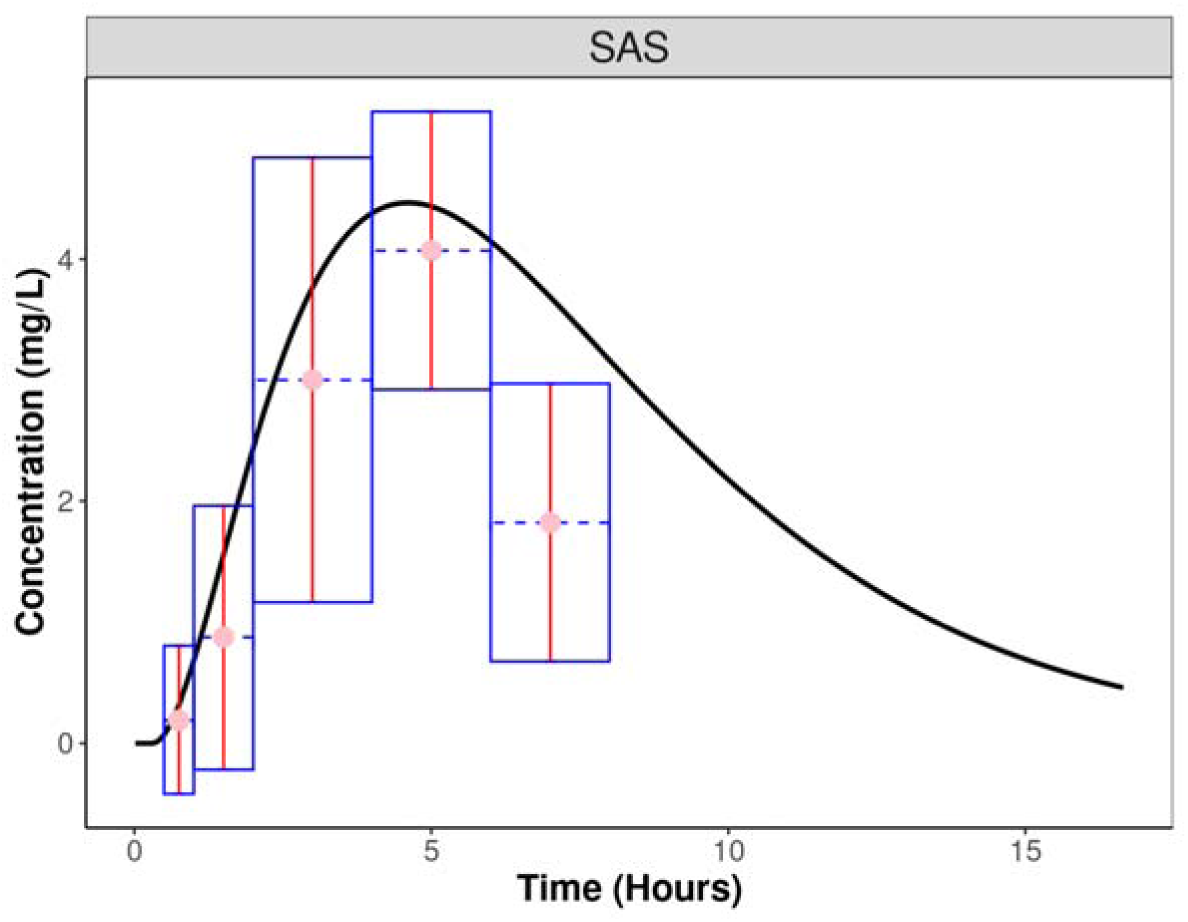
Comparison of predicted and observed moxifloxacin concentrations in the subarachnoid space (SAS) of human subjects without meningitis. Model predictions are shown as solid black line. Observed human data (21) were represented as pink circles, with vertical (red) and horizontal (blue dashed) error bars indicating standard deviations and sampling-time intervals, respectively.

### 3.3 Model Validation in TBM Patients

The adapted TBM model successfully reproduced observed plasma and SAS concentration-time profiles from multiple clinical studies (Figure 4). Predicted plasma AUC_0-24_, C_max_, and t□/□ values were consistent with observed medians for both 400□mg and 800□mg dosing scenarios (Figure 4A). While the predicted AUC_0-24_ and C_max_ values increased proportionally with dose, the observed data indicated a degree of nonlinearity between the two dosing levels (4). Across TBM studies, most observations, except for lower values reported from one case study (24), fell within ±2-fold of the model predictions (Figure 4B), suggesting a good model predictive performance.

**Figure 4:**
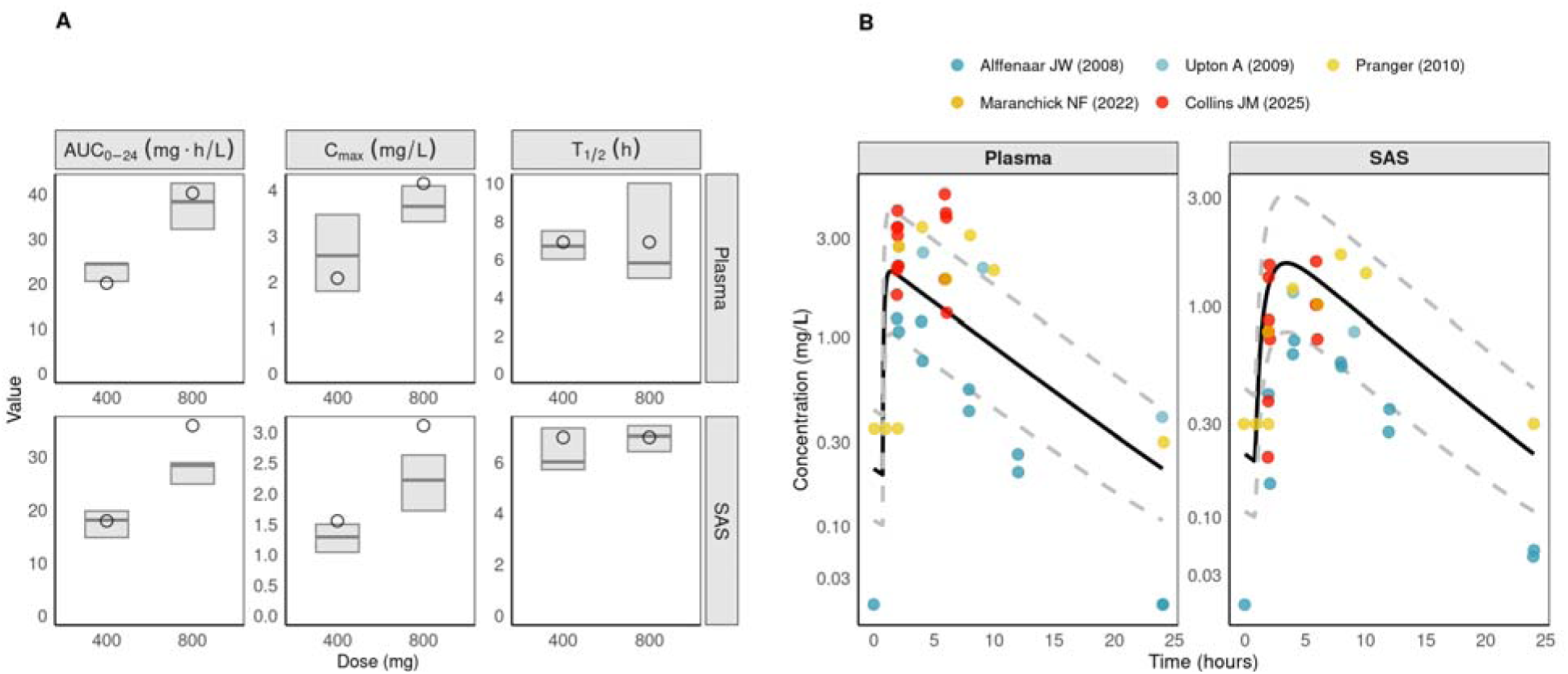
Validation of human CNS PBPK model predictions for moxifloxacin in TBM patients co-administered with rifampicin. A. Boxplots of model-predicted pharmacokinetic parameters—area under the concentration–time curve from 0 to 24 hours (AUC_0-24_), maximum concentration (C_max_), and half-life (t_1/2_)—in plasma (top row) and subarachnoid space (SAS; bottom row) at 400□mg and 800□mg doses. Observed reference values (4) are depicted as horizontal lines (median) and shading (interquartile range, 25th–75th percentile) within each boxplot; predicted values are shown as circles. B. Model-predicted typical concentration–time profiles (black lines) in plasma and SAS compared with observed clinical data (colored dots) from multiple TBM studies (14, 15, 22–24). Dashed grey lines represent a two-fold error range around the model-predicted curves and the y axis is logarithmic. The observed concentrations of both plasma and SAS were considerably lower from one case report (24) than the other TBM studies.

### 3.4 Comparative PK/PD Analysis of Moxifloxacin in Plasma and Key CNS Compartments

Simulated steady-state PK profiles revealed regional heterogeneity in moxifloxacin exposure across CNS compartments (Figure 5A). Concentrations were highest in the SAS, followed by the CM and brain VENs, and lowest in the brain ECF. Coadministration of rifampicin reduced exposures in plasma and all CNS compartments. Regarding the drug efficacy evaluation using an fAUC_0-24_/MIC threshold of 53 (Figure 5B), the 400 and 600 mg once-daily regimens did not achieve the target in most CNS compartments, whereas 800 and 1000 mg generally did when moxifloxacin was administered alone. In combination with rifampicin, only the 1000 mg regimen attained the target in most compartments except for brain ECF that remained sub-target (fAUC_0-24_/MIC ≈ 42).

**Figure 5:**
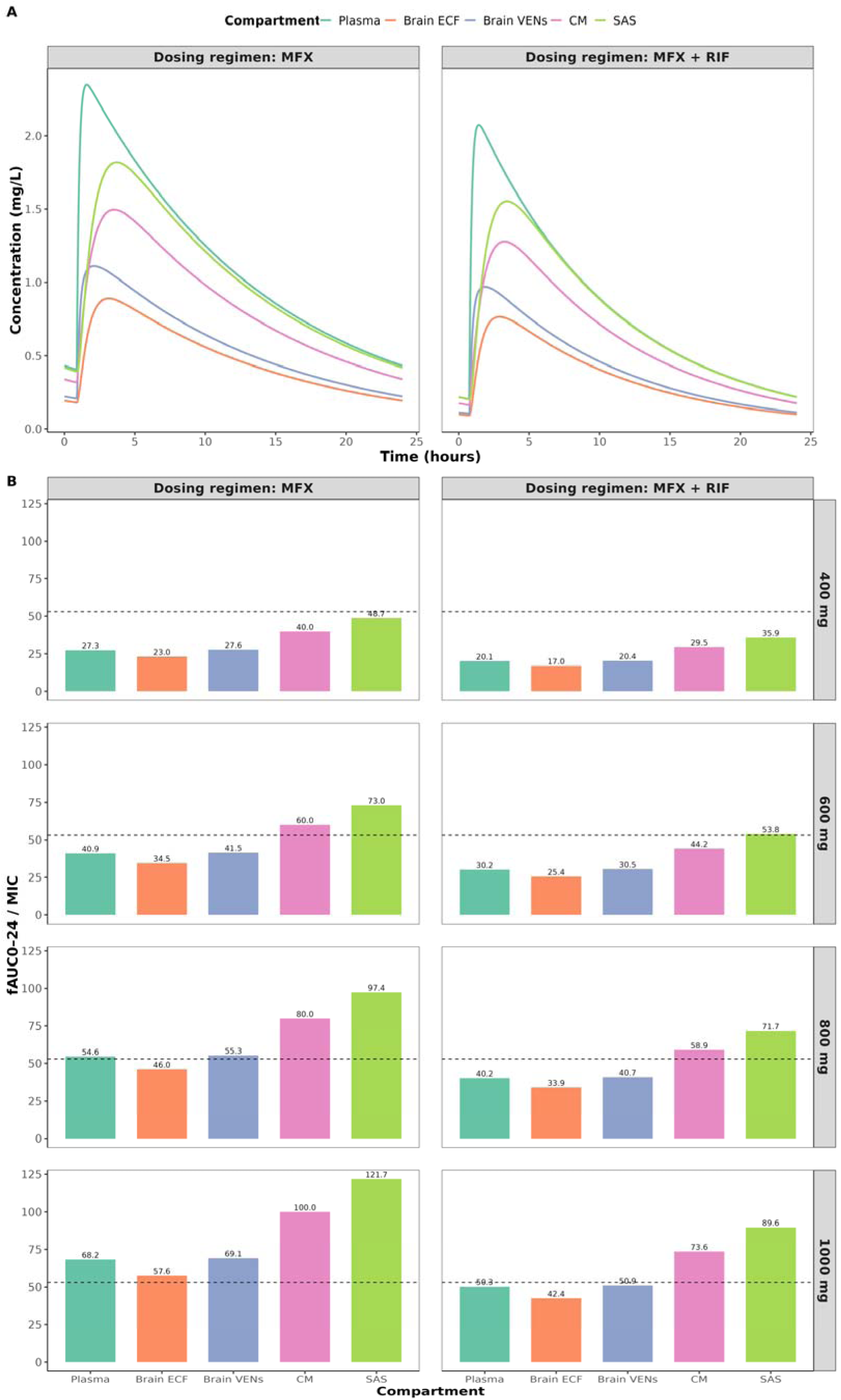
Moxifloxacin (MFX) pharmacokinetic (PK) profiles and target attainment with and without rifampicin (RIF). A. Simulated steady-state concentration-time profiles of MFX under standard dosing (400 mg oral once daily) in plasma and key CNS compartments: brain extracellular fluid (Brain ECF), brain ventricles (Brain VENs), cisterna magna (CM), and subarachnoid space (SAS). Left: MFX alone; right: MFX + RIF. Colors denoted compartments. B. Bar plots of the ratio of the 24-hour area under the concentration–time to the minimal inhibitory concentration (fAUC_0–24_/MIC) based on 400, 600, 800, and 1000 mg once per day of MFX (rows) with or without RIF (columns). The dashed horizontal line at 53 marked the fAUC_0-24_/MIC target (36).

## 4 Discussion

Current understanding of moxifloxacin PK in the human CNS remains limited to sparse CSF data, providing little insight into regional exposure or the impact of rifampicin co-administration. This study addressed these gaps by quantitatively describing moxifloxacin CNS distribution leveraging a cross-species translational PBPK modeling approach. Simulations showed regional concentration differences in the CNS compartments (SAS > CM > brain VENs > brain ECF). Co-administration with rifampicin substantially reduced moxifloxacin exposure in plasma and, consequently, across CNS compartments. Relative to the PK/PD target fAUC□–□□/MIC of 53, higher moxifloxacin doses were predicted to improve target attainment in key CNS sites particularly when combined with rifampicin in TBM.

We found that estimated moxifloxacin drug concentrations differed significantly across human CNS compartments. To date, human moxifloxacin CNS samples have only been obtained from SAS. For comparison, only a few other drugs have been systematically studied for their CNS PK exposure in humans, and the corresponding data align with our simulated findings of CNS regional heterogeneity (40–43). Even lipophilic agents such as linezolid and metronidazole were not homogenously distributed within the CNS, with higher concentrations in brain VENs than in brain ECF (40, 41), while the SAS was consistently the compartment with highest concentrations (42, 43). Our simulations also indicated high BCSFB penetration of moxifloxacin with an unbound AUC ratio (brain VENs/plasma) approaching 1. Similar values have been observed for ofloxacin, which has similar chemical structure with moxifloxacin, under both inflamed and non-inflamed meningeal conditions (44, 45). Therefore, plasma or lumbar CSF samples may overestimate therapeutic coverage in other sites such as brain ECF.

This regional heterogeneity in CNS drug exposure has direct implications for dose optimization, as therapeutic success in TBM depends mainly on achieving adequate drug levels at the relevant CNS sites of infection. In a randomized trial in TBM with limited patients size (n=19), regimens including higher-dose moxifloxacin (800 mg) did not improve clinical outcomes versus standard 400 mg (46). Our simulation results provide a mechanistic insight: under rifampicin co-administration, CNS exposures fall proportionally with the systemic decrease, leaving several infected regions below PK/PD target at 400-800 mg. Rifampicin, a broad inducer of drug-metabolizing enzymes/transporters (47), reduced moxifloxacin systemic AUC_0-24_ by ∼31% (12) while preserving dose-proportional PK in plasma and CSF from 400 to 800 mg (46). We assumed rifampicin’s impact on moxifloxacin’s PK as a systemic effect, since there is no human evidence that rifampicin meaningfully alters moxifloxacin CNS PK via local induction on enzymes or transporters at the BBB/BCSFB/brain. The expression levels of uridine diphosphate glucuronosyltransferase and sulphotransferase, which play important roles in moxifloxacin disposition (48), are markedly lower in the CNS barriers compared to that in liver (49, 50). Thus, rifampicin likely has a minimal or no effect on CNS-specific disposition of moxifloxacin, and CNS exposures fall largely in proportion to the systemic decrease. Our results emphasize reassessing moxifloxacin dosing strategies in rifampicin-containing TBM regimens, where reduced CNS exposure may compromise efficacy especially in patients with brain ECF involvement. Nonetheless, while the results suggested that doses > 1000 mg may be required to meet moxifloxacin PK/PD target fAUC□–□□/MIC > 53 across all infected CNS sites during rifampicin concomitant treatment, extrapolation above 800 mg should be cautious because non-linearities have been observed from 400 to 800 mg in TBM, e.g., CSF:plasma AUC ratios decreased at 800 mg in clinical sampling (4). Additionally, the current study focused on the target attainment of moxifloxacin alone, while rifampicin also contributes bactericidal activity within the CNS. Rifampicin’s direct effect on bacterial killing or and potential pharmacodynamic interactions between the two agents warrant further evaluation, but these aspects could not be assessed in the current study (69). Meanwhile, a large-scale clinical study has been initiated to continue assessing the therapeutic value of intensified rifampicin and moxifloxacin dosing in TBM (51). As TBM pathogenesis likely progresses from brain ECF to ventricular CSF and eventually to the SAS (52), maintaining adequate drug levels throughout this pathway is critical. Disease severity may shift bacillary burden toward the SAS in later stages (17), suggesting the need to map stage-dependent target sites to optimize dosing.

Our model was designed to mechanistically characterize moxifloxacin disposition in the CNS while representing systemic PK by a one-compartment model. For trans-barrier drug movement across the BBB/BCSFB, we directly estimated the drug permeability rates from the pig concentration-time data, rather than partitioning into passive and active components due to limited mechanistic data existing for active moxifloxacin transport at the BBB. Regarding the CSF turnover, classical CNS PBPK models (53, 54) held the historical assumption (55) that constant, unidirectional CSF circulation equal to choroid-plexus secretion rate across all CSF spaces and returning from SAS to the blood. We instead implemented bidirectional CSF dynamics between CM and SAS with opposing flows, QCSF_DIAS_ (towards head) and QCSF_SYS_ (away from head), better reflecting MRI-observed bidirectional CSF dynamics (56, 57) and therefore providing a more physiologically accurate representation of CNS drug transport (58). We also estimated a unidirectional CSF reabsorption rate from SAS into plasma rather than fixing it to choroid-plexus production rate since the current evidence (59–61) suggested choroid-plexus secretion alone does not account for total CSF turnover. Given no direct human measurements for the CNS-specific parameters (QECF, QCSF_VENs_, QCSF_DIAS_, QCSF_SYS_, QCSF_REB_), we used CNS-relevant interspecies scaling, including human-to-pig surface area ratios for BBB and BCSFB permeability parameter and brain volume ratio for CNS flow rates. The classical allometry (Kleiber’s law) uses the body mass ratio as scaling base and is commonly used to predict the systemic parameter such as plasma clearance rate, but might not be suitable for CNS specific parameters (62). Our scaling approach showed better predictive accuracy (Figure S5) compared to the classical scaling, suggesting that CNS-relevant scaling ratios may be prioritized over default body mass when translating CNS parameters. Because mammalian brain density averages ∼1 g/mL, brain volume and weight could be effectively interchangeable as scaling bases in the practice (63–65). Moreover, higher exponent of 0.95 for the CNS parameter scaling might be needed since the classical 0.75 exponent represents basal metabolic rate and may not reflect relatively high brain metabolic demands (66).

It is worth noting that the presented modeling approach did not incorporate the mechanisms such as enzyme or transporter-mediated drug disposition that could cause non-linear PK and thus exhibited dose-proportional (linear) moxifloxacin kinetics. This assumption appears reasonable, as clinical data reported linear PK across the tested range of at least 50–800 mg (67). The model further assumed uniform CNS physiology and BBB/BCSFB barrier integrity since moxifloxacin CNS penetration is generally favorable even under non-inflamed conditions (21). However, TBM may produce highly variable and localized inflammation that affects drug penetration and limits CSF flow rates in case of obstructive hydrocephalus (68), highlighting the need for additional data to quantify these effects. Moreover, clinical validation relied on sparse and heterogeneous PK data from case reports and small cohorts, which limits the precision of model verification. Incorporating patient-level variability, disease staging, and longitudinal data would strengthen predictive accuracy. Comprehensive CNS PK data are also required to perform PK/PD analyses, especially to calculate the *f*AUC_0-24_/MIC, the key predictor of fluoroquinolone efficacy in CNS infections. Finally, It should be noted that the commonly used *f*AUC_0-24_/MIC threshold of 53 was derived using the plasma PK data (36), while there is currently no validated CNS-specific cutoff exists for TBM. Deriving CNS-specific PK/PD target is therefore warranted for the future efforts, and possibly linking PK/PD to clinical outcomes as well. (69)

## Conclusion

In conclusion, we used a translational cross-species PBPK modeling approach to characterize regional CNS moxifloxacin exposure and to inform optimal dosing strategies. We found that higher dose might be needed to ensure adequate therapeutic coverage in TBM when moxifloxacin is combined with rifampicin. Our findings underscore the need to consider regional CNS differences and drug-drug interactions particularly with rifampicin when designing effective treatment strategies.

## Data Availability

All data produced in the present study are available upon reasonable request to the authors

## Supplementary document

**Table S1.** Study characteristics of moxifloxacin pharmacokinetics in tuberculosis meningitis.

| Study Name | NO. Patients | Dose (Frequency, Administration Route) | Coadministered with rifampicin | Study Design | Sampling Compartment | TBM Severity |
| --- | --- | --- | --- | --- | --- | --- |
| Collins et al., 2025 | 9 | Moxifloxacin (400 mg once daily, oral) | Yes | Patients receiving drug treatment for $\geq 7$ days; samples taken 2 h and 6 h post-dose at steady state | Plasma & CSF | Mixed diagnostic categories: 29% confirmed vs. 71% probable TBM; low to moderate severity grade I or II based on Glasgow coma scale. |
| Alffenaar et al., 2008 | 1 | Moxifloxacin (400 mg or 800 mg once daily, oral) | Yes | Steady-state PK profiles measured at multiple time points on day 3 after each dose. | Plasma & CSF | Nonactive TBM. |
| Alffenaar et al., 2009 | 4 | Moxifloxacin (400 mg or 800 mg once daily, oral) | Yes | Steady-state concentration–time profiles obtained for each dose while only PK parameters $AUC_{0-24}$, $C_{max}$ and $T_{1/2}$ were reported per dose. | Plasma & CSF | Active TBM without clearly stating the severity grade. |
| Pranger et al., 2010 | 1 | Moxifloxacin (400 mg once daily, oral) | Not reported | Steady-state PK profiles measured at multiple time points. | Plasma & CSF | Not reported |
| Upton et al., 2009 | 1 | Moxifloxacin (400 mg once | Yes | Steady-state PK profiles measured | Plasma & CSF | Non-severe case due to no |
|  |  | daily, oral) |  | at multiple time points. |  | Mtb was seen after the previous treatment. |
| Maranchick et al., 2022 | 10 | Moxifloxacin (400 mg once daily, oral) | Yes | Patients receiving drug treatment for $\geq 7$ days; samples taken 2 h and 6 h post-dose at steady state | Plasma & CSF | 8% confirmed TBM; Mild to moderate symptom based on glasgow coma scale (9-15). |

**Table S2.** Physiological parameters (fixed value in the model)

| Systemic parameter |  | Fixed in pig model |  | Fixed in human model |  |
| --- | --- | --- | --- | --- | --- |
|  |  | Value | Reference | Value | Reference |
| Volumes (mL) | Total brain ($V_{tot}$) | 107 | (70): 103 mL, female pigs.<br>(71) :107 mL, male pigs.<br>(72): 108 mL, adult landrace pigs. | 1387 | Average value for male is 1442, and for female is 1332 (73). |
| | Brain intracellular fluid ($V_{icf}$) | 85.6 | (74): Intracellular brain fluid volume is estimated to be approximately 80% of the total brain volume ($107 \times 80\% = 85.6$ mL). | 1109.6 | Brain ICF volume was assumed 80% of total brain volume (75). |
| | Brain extracellular fluid ($V_{ecf}$) | 21.4 | (74): ECF volume fraction of $V_{tot}$ is reported as value around 20% ($107 \times 20\% = 21.4$ mL). | 277.4 | Brain ECF volume was assumed 20% of total brain volume (75). |
| | Total cerebrospinal fluid ($V_{csf}$) | 19.7 | (71): 19.55mL male pigs.<br>(76): 20mL, male pigs. | 292 | 254-331 mL of CSF was distributed across cranial and spinal space (77). |
| | Brain ventricles ($V_{BVEN}$) | 2.4 | male pig, 1.98 mL of lateral ventricles (78); piglets, 0.4 mL of 3<sup>rd</sup> & 4<sup>th</sup> ventricles (79). | 26 | Around 26mL fills the ventricles, including 23mL of lateral ventricles and 3mL of 3<sup>rd</sup> & 4<sup>th</sup> ventricles (80). |
| | Cisterna magna ($V_{CM}$) | 1 | (81, 82): no direct evidence, but only 1 mL of CSF could be safely withdrawn from the cisterna magna. | 6.5 | 3-10 mL (83). |
| | Subarachnoid space ($V_{SAS}$) | 16.3 | This volume was calculated by subtracting the volume of the remaining CSF regions from the total volume. | 260 | This volume was calculated by subtracting the volume of the remaining CSF regions from the total volume. |

**Table S3.**
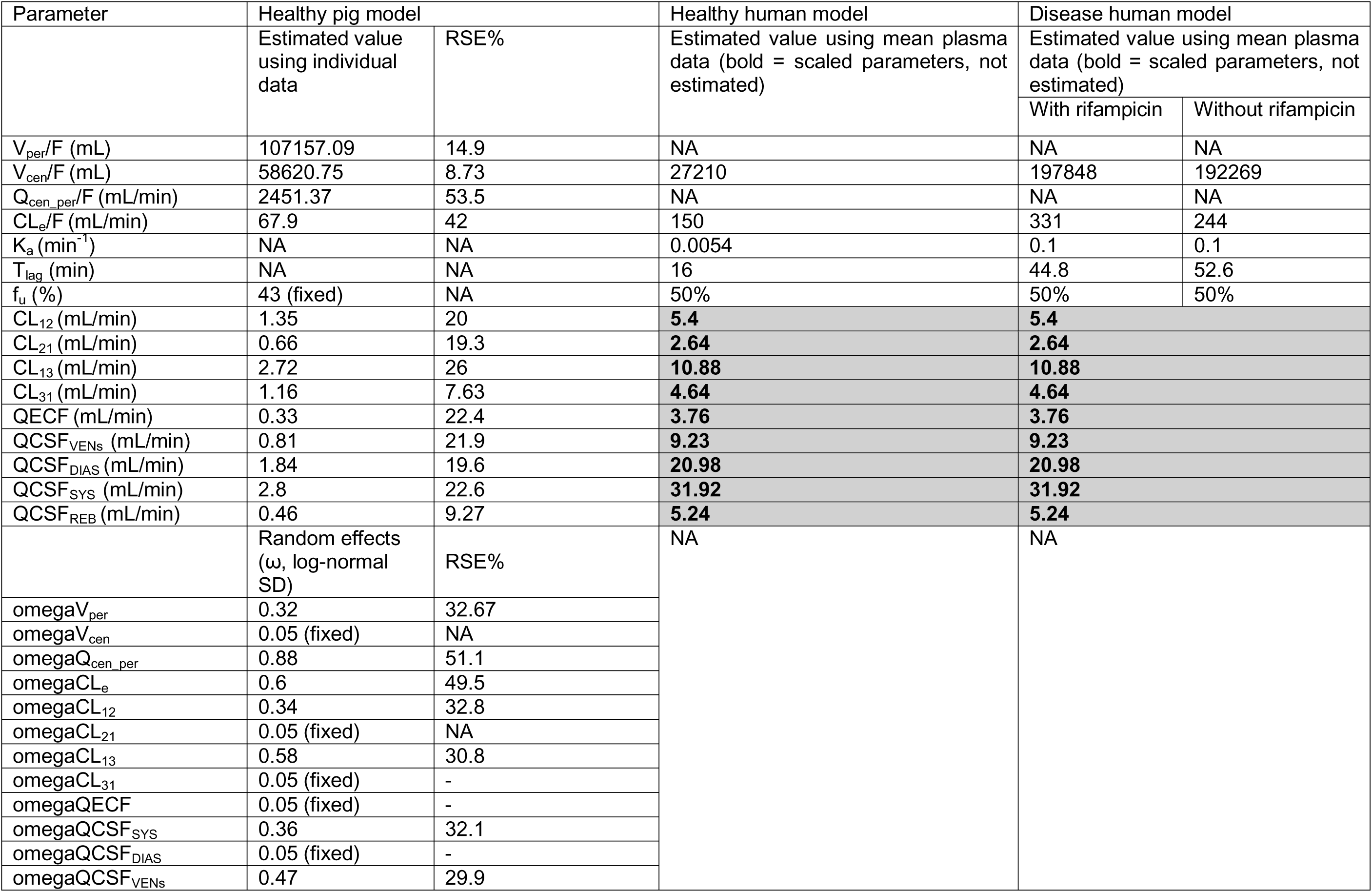
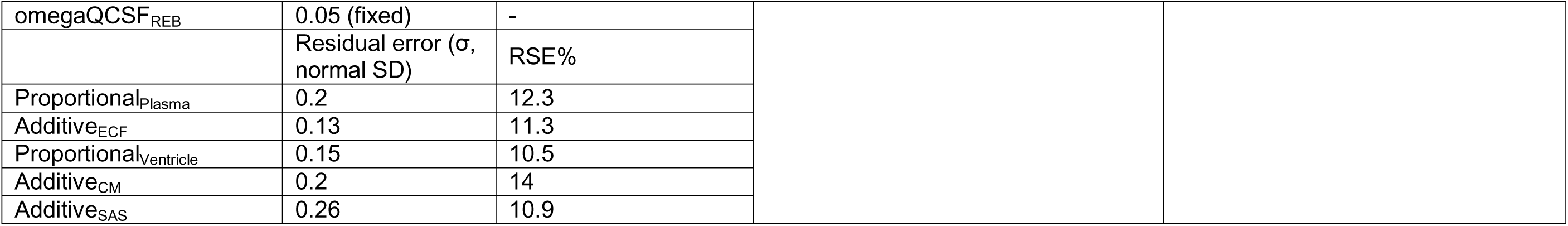
The remaining input parameters of moxifloxacin in healthy pig, healthy and disease human models.

**Table S4.** Scaling methods used for translating related parameters from pig to human in healthy conditions.

|  | Equations |  |  |  |  |  |  |  |  |  |
| --- | --- | --- | --- | --- | --- | --- | --- | --- | --- | --- |
| | $P_h = P_p \times \frac{BW_h^{0.75}}{BW_p}$ | | $P_h = P_p \times \frac{V_{brh}^{0.67}}{V_{brp}}$ | | $P_h = P_p \times \frac{V_{brh}^{0.84-1.06}}{V_{brp}}$ | | $P_h = P_p \times \frac{W_{brh}^{0.67}}{W_{brp}}$ | | $P_h = P_p \times \frac{SA_{BBB/BCSFB,h}}{SA_{BBB/BCSFB,p}}$ | |
| Parameter value | BW<sub>h</sub> | 70 kg<sup>a</sup> | Vbr<sub>h</sub> | 1387 mL<sup>b</sup> | Vbr<sub>h</sub> | 1387 mL<sup>b</sup> | Wbr<sub>h</sub> | 1350 g (84) | SABBB<sub>h</sub> | 10m<sup>2</sup>(85)<sup>c</sup> |
|  |  |  |  |  |  |  |  |  | SABCSFB<sub>h</sub> | 5m<sup>2</sup>(85)<sup>d</sup> |
|  | BW<sub>p</sub> | 40 kg<sup>a</sup> | Vbr<sub>p</sub> | 107 mL<sup>b</sup> | Vbr<sub>p</sub> | 107 mL<sup>b</sup> | Wbr<sub>p</sub> | 110 g (86) | SABBB<sub>p</sub> | 2.5m<sup>2</sup>(87)<sup>e</sup> |
|  |  |  |  |  |  |  |  |  | SABCSFB<sub>p</sub> | 1.25m<sup>2</sup>(85)<sup>d</sup> |
| Notes | This method uses a 0.75 exponent based on body weight (BW). This classic allometric scaling factor is often used to translate CNS parameters from rodents to humans (19). |  | This method employs a 0.67 exponent based on brain volume (Vbr). This reflects scaling of permeability surface areas and structural proportions in CNS translation (42). |  | This method uses an exponent range of 0.84–1.06 (0.95 used) based on brain volume (Vbr), capturing metabolic and hemodynamic factors observed in cross-species cortical blood flow scaling (28). |  | This method applies a 0.67 exponent based on brain weight (Wbr), assuming similar tissue architecture and enabling tissue permeability scaling from rat to human CNS parameters (88). |  | This method has commonly been used to scaling the drug permeability across the membrane from in vitro cell line to human. |  |
| Application | Barrier permeability | CSF flows | Barrier permeability |  | CSF flows |  | Barrier permeability |  | Barrier permeability |  |
| Method 1 |  |  |  |  | X |  |  |  | X |  |
| Method 2 |  |  | X |  | X |  |  |  |  |  |
| Method 3 |  |  |  |  | X |  | X |  |  |  |
| Method 4 | X | X |  |  |  |  |  |  |  |  |

<sup>a</sup> Typical values for human and minipigs.
<sup>b</sup> Values are the same as those in Table S1, with corresponding references.
<sup>c</sup> Estimated range from 12-17 m<sup>2</sup> (14 m<sup>2</sup> was used).
‡ Assumed to be 50% of BBB.
‡ Based on capillary surface area of newborn pig cortex, with a value of 23 mm<sup>2</sup>/mm<sup>3</sup>.

**Figure S1:**
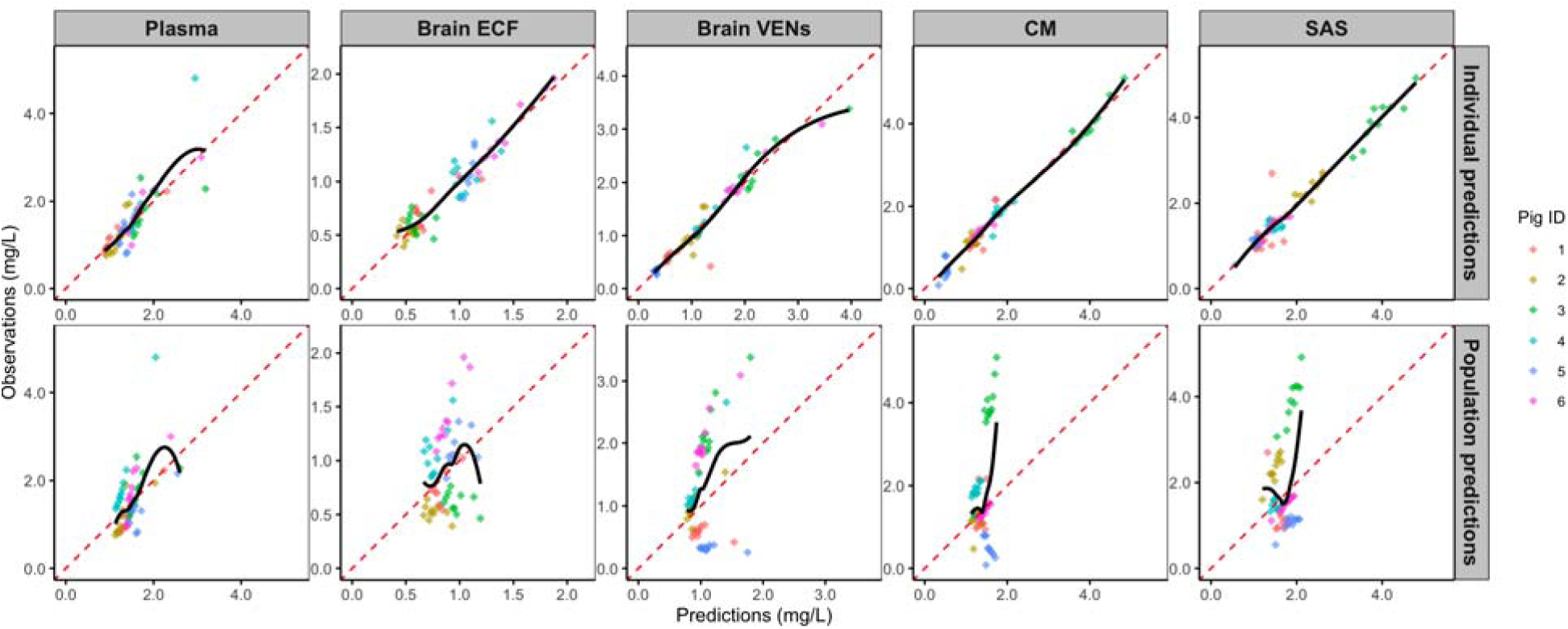
Goodness-of-fit analysis for individual and population predictions. Goodness-of-fit plots comparing observed versus predicted moxifloxacin concentrations for individual (top row) and population (bottom row) predictions in plasma, brain extracellular fluid (ECF), brain ventricles (VENs), cisterna magna (CM), and subarachnoid space (SAS). The red dashed lines represent the line of unity, indicating perfect agreement between observed and predicted values. And the spline shows the trend in predictions.

**Figure S2:**
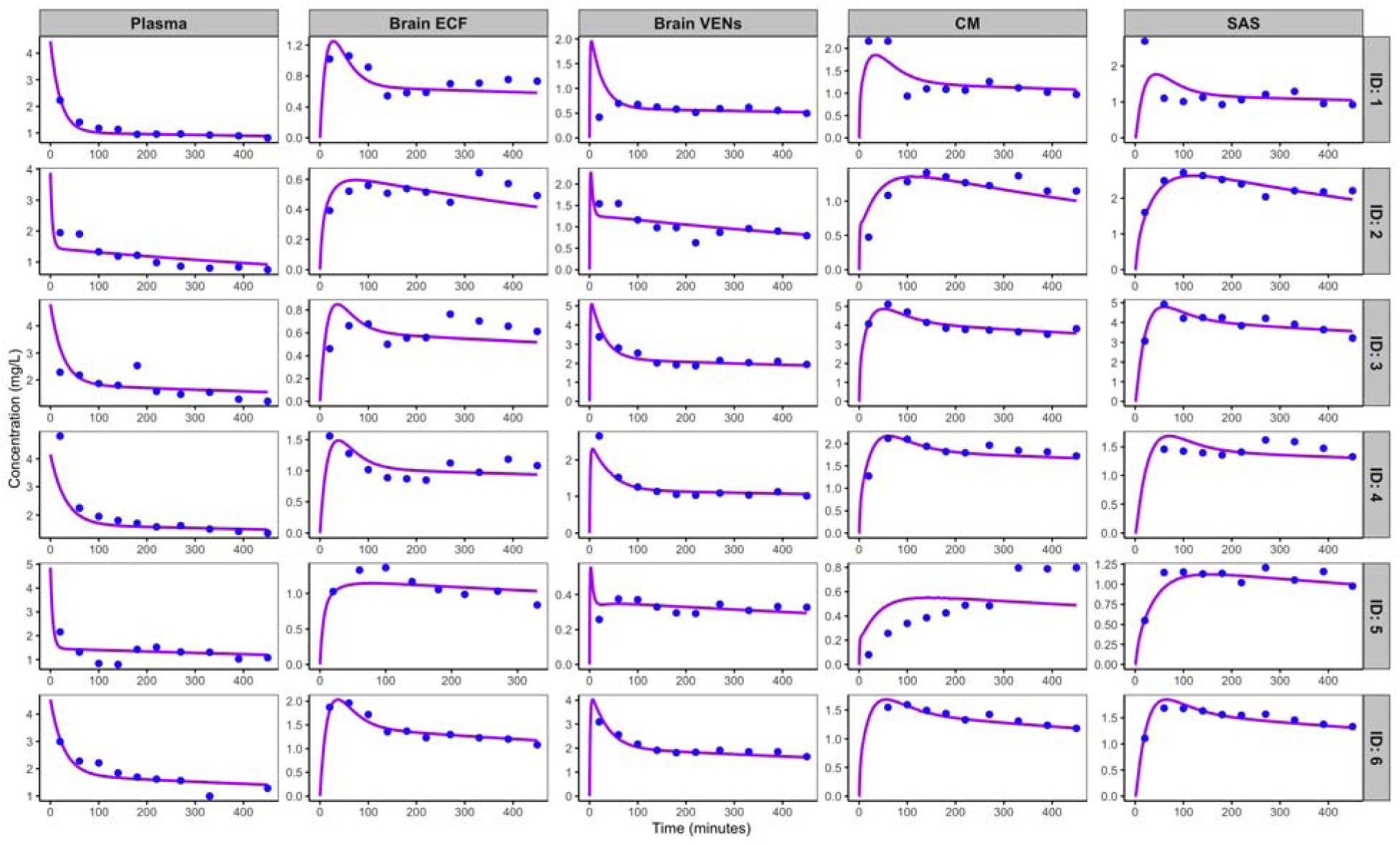
Individual model fitting of moxifloxacin concentration-time profiles. Model fit for each individual pig (ID 1–6) across different compartments [plasma, brain extracellular fluid (ECF), brain ventricles (VENs), cisterna magna (CM), and subarachnoid space (SAS)]. Blue circles indicate observed data points, while the magenta lines represent the model-predicted concentration-time profiles for each individual.

**Figure S3:**
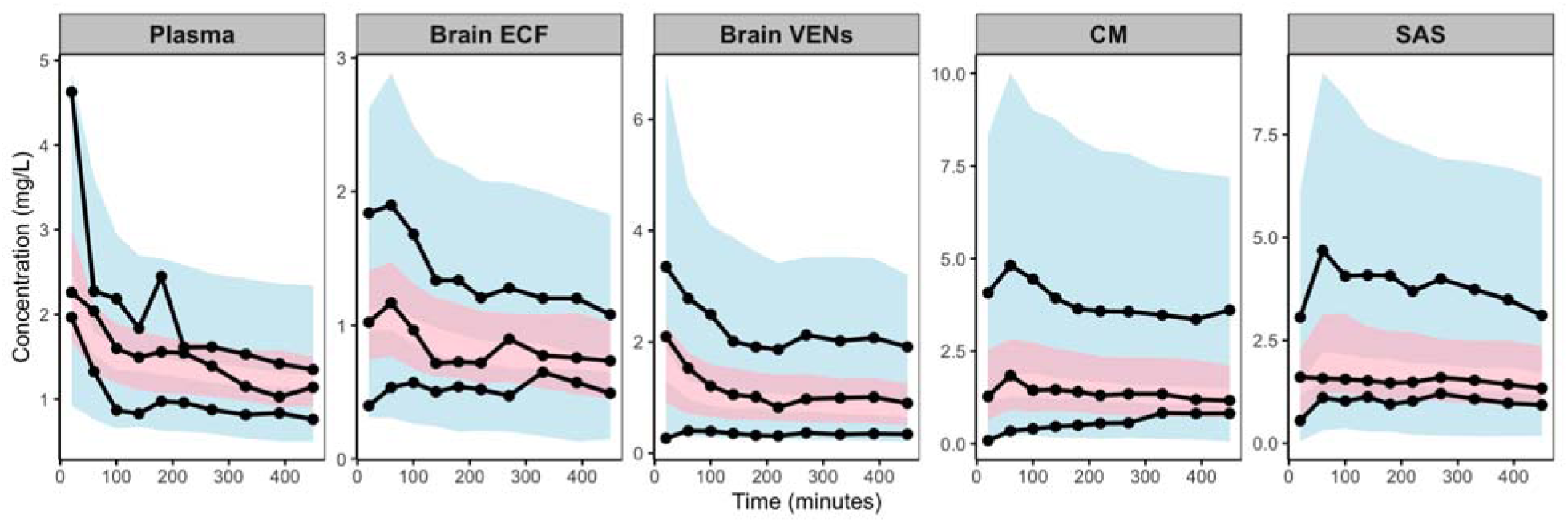
Visual predictive check (VPC) of moxifloxacin concentrations. Visual predictive check (VPC) for moxifloxacin concentrations in plasma, brain extracellular fluid (ECF), brain ventricles (VENs), cisterna magna (CM), and subarachnoid space (SAS). Solid lines represent the median and 90% prediction intervals of the model, while the shaded areas correspond to the 90% confidence intervals of the predicted data. The black dots represent observed data from individual pigs.

**Figure S4:**
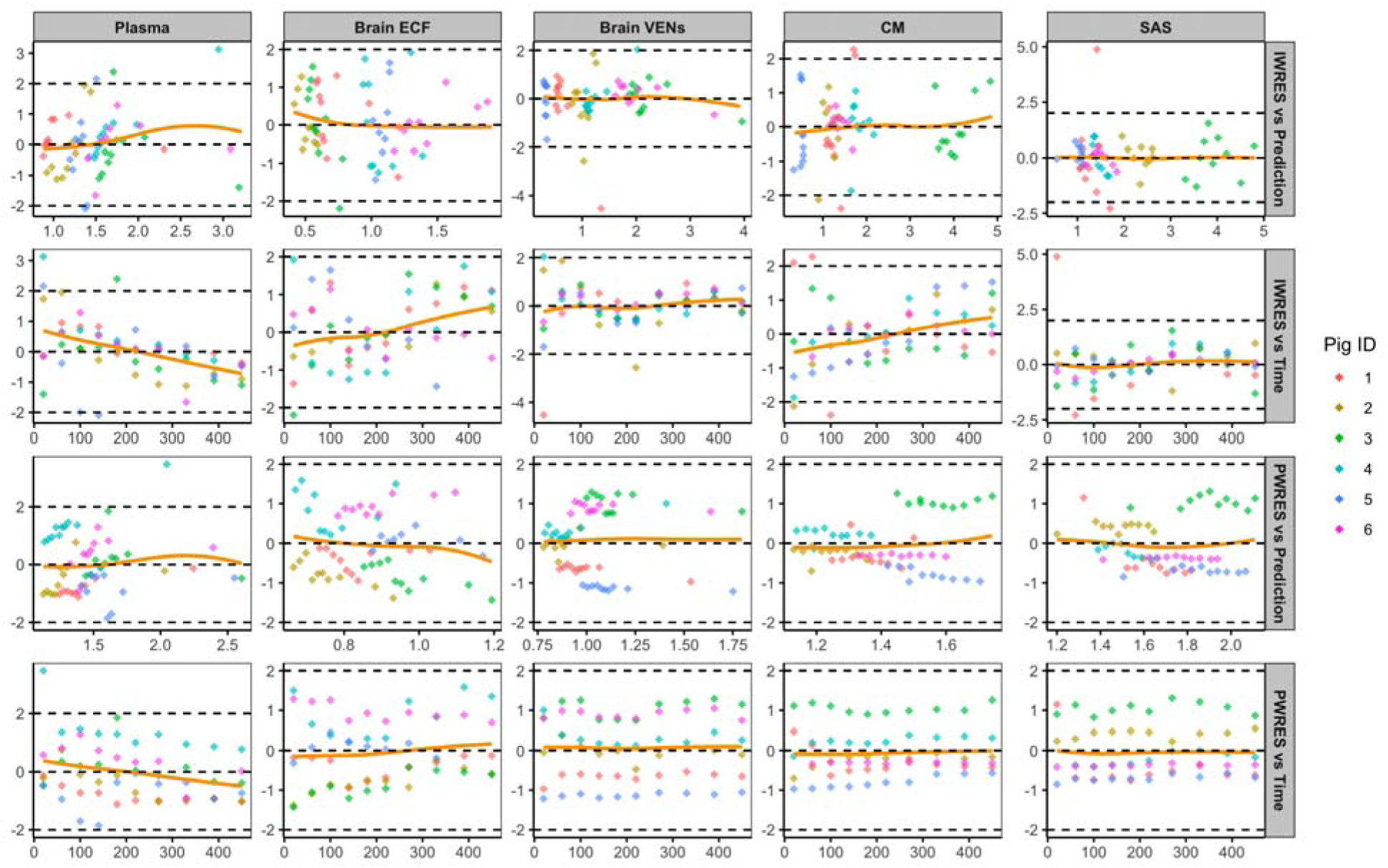
Residual analysis: IWRES and PWRES plots. Individual weighted residuals (IWRES) versus model predictions and time (Upper two panels) and population weighted residuals (PWRES) versus model predictions and time (Lower two panels) for moxifloxacin concentrations in plasma, brain extracellular fluid (ECF), brain ventricles (VENs), cisterna magna (CM), and subarachnoid space (SAS). The dashed lines represent the 95% confidence limits for the residuals, and the orange line shows the trend in the residual distribution.

**Figure S5:**
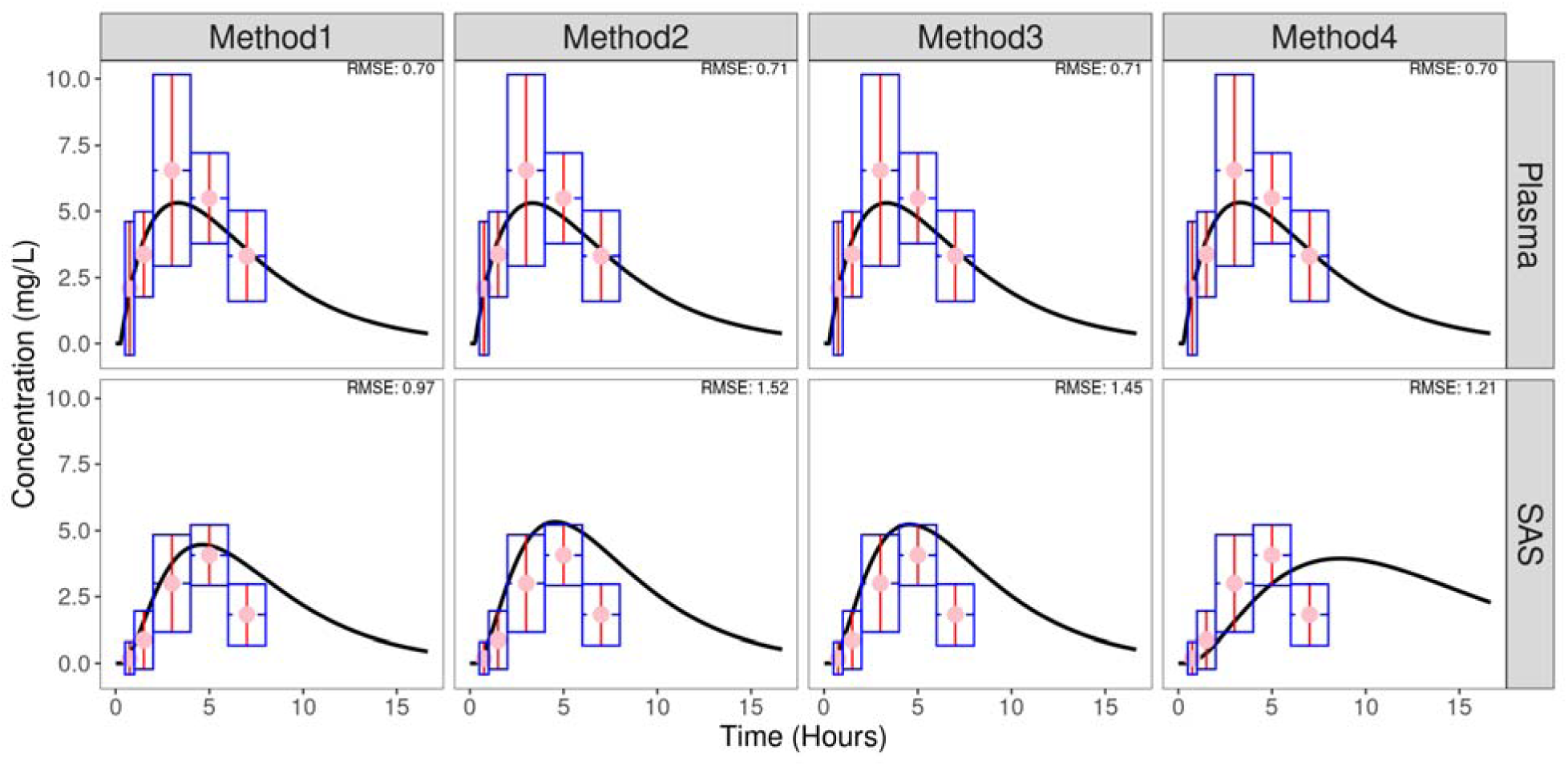
Comparison of predicted and observed moxifloxacin concentrations in the plasma and subarachnoid space (SAS) of human subjects without meningitis using four allometric scaling methods (Methods 1–4). Method 1-4 predictions are shown as solid black lines. Observed human data are represented as pink circles, with vertical (red) and horizontal (blue dashed) error bars indicating standard deviations and sampling-time intervals, respectively. Root mean square error (RMSE) values, quantifying model prediction accuracy, are provided for each method.

## Reference

1. Stadelman AM, Ellis J, Samuels THA, Mutengesa E, Dobbin J, Ssebambulidde K, et al. Treatment Outcomes in Adult Tuberculous Meningitis: A Systematic Review and Meta-analysis. Open Forum Infect Dis. 2020;7(8):ofaa257.

2. Chiang SS, Khan FA, Milstein MB, Tolman AW, Benedetti A, Starke JR, et al. Treatment outcomes of childhood tuberculous meningitis: a systematic review and meta-analysis. Lancet Infect Dis. 2014;14(10):947–57.

3. Török ME. Tuberculous meningitis: advances in diagnosis and treatment. Br Med Bull. 2015;113(1):117–31.

4. Alffenaar JWC, van Altena R, Bökkerink HJ, Luijckx GJ, van Soolingen D, Aarnoutse RE, et al. Pharmacokinetics of moxifloxacin in cerebrospinal fluid and plasma in patients with tuberculous meningitis. Clin Infect Dis. 2009;49(7):1080–2.

5. Heemskerk AD, Bang ND, Mai NT, Chau TT, Phu NH, Loc PP, et al. Intensified Antituberculosis Therapy in Adults with Tuberculous Meningitis. N Engl J Med. 2016;374(2):124–34.

6. Davis AG, Wasserman S, Stek C, Maxebengula M, Jason Liang C, Stegmann S, et al. A Phase 2A Trial of the Safety and Tolerability of Increased Dose Rifampicin and Adjunctive Linezolid, With or Without Aspirin, for Human Immunodeficiency Virus–Associated Tuberculous Meningitis: The LASER-TBM Trial. Clinical Infectious Diseases. 2022;76(8):1412–22.

7. Rodríguez JC, Ruiz M, López M, Royo G. In vitro activity of moxifloxacin, levofloxacin, gatifloxacin and linezolid against Mycobacterium tuberculosis. Int J Antimicrob Agents. 2002;20(6):464–7.

8. Nuermberger EL, Yoshimatsu T, Tyagi S, O’Brien RJ, Vernon AN, Chaisson RE, et al. Moxifloxacin-containing regimen greatly reduces time to culture conversion in murine tuberculosis. Am J Respir Crit Care Med. 2004;169(3):421–6.

9. Nau R, Sörgel F, Eiffert H. Penetration of drugs through the blood-cerebrospinal fluid/blood-brain barrier for treatment of central nervous system infections. Clin Microbiol Rev. 2010;23(4):858–83.

10. WHO Guidelines Approved by the Guidelines Review Committee. WHO consolidated guidelines on drug-resistant tuberculosis treatment. Geneva: World Health Organization © World Health Organization 2019.; 2019.

11. Dorman SE, Nahid P, Kurbatova EV, Phillips PPJ, Bryant K, Dooley KE, et al. Four-Month Rifapentine Regimens with or without Moxifloxacin for Tuberculosis. New England Journal of Medicine. 2021;384(18):1705–18.

12. Nijland HM, Ruslami R, Suroto AJ, Burger DM, Alisjahbana B, van Crevel R, et al. Rifampicin reduces plasma concentrations of moxifloxacin in patients with tuberculosis. Clin Infect Dis. 2007;45(8):1001–7.

13. Donald PR. Cerebrospinal fluid concentrations of antituberculosis agents in adults and children. Tuberculosis (Edinburgh, Scotland). 2010;90(5):279–92.

14. Collins JM, Kipiani M, Jin Y, Sharma AA, Tomalka JA, Avaliani T, et al. Pharmacometabolomics in TB meningitis-Understanding the pharmacokinetic, metabolic, and immune factors associated with anti-TB drug concentrations in cerebrospinal fluid. PloS One. 2025;20(3):e0315999.

15. Maranchick NF, Alshaer MH, Smith AGC, Avaliani T, Gujabidze M, Bakuradze T, et al. Cerebrospinal fluid concentrations of fluoroquinolones and carbapenems in tuberculosis meningitis. Frontiers In Pharmacology. 2022;13:1048653.

16. Wilkinson RJ, Rohlwink U, Misra UK, van Crevel R, Mai NTH, Dooley KE, et al. Tuberculous meningitis. Nat Rev Neurol. 2017;13(10):581–98.

17. Be NA, Kim KS, Bishai WR, Jain SK. Pathogenesis of central nervous system tuberculosis. Curr Mol Med. 2009;9(2):94–9.

18. Gerlowski LE, Jain RK. Physiologically based pharmacokinetic modeling: principles and applications. J Pharm Sci. 1983;72(10):1103–27.

19. Yamamoto Y, Välitalo PA, Wong YC, Huntjens DR, Proost JH, Vermeulen A, et al. Prediction of human CNS pharmacokinetics using a physiologically-based pharmacokinetic modeling approach. Eur J Pharm Sci. 2018;112:168–79.

20. Mariager T, Terkelsen JH, Bue M, Öbrink-Hansen K, Nau R, Bjarkam CR, et al. Continuous evaluation of single-dose moxifloxacin concentrations in brain extracellular fluid, cerebrospinal fluid, and plasma: a novel porcine model. J Antimicrob Chemother. 2024;79(6):1313–9.

21. Kanellakopoulou K, Pagoulatou A, Stroumpoulis K, Vafiadou M, Kranidioti H, Giamarellou H, et al. Pharmacokinetics of moxifloxacin in non-inflamed cerebrospinal fluid of humans: implication for a bactericidal effect. J Antimicrob Chemother. 2008;61(6):1328–31.

22. Upton A, Woodhouse A, Vaughan R, Newton S, Ellis-Pegler R. Evolution of central nervous system multidrug-resistant Mycobacterium tuberculosis and late relapse of cryptic prosthetic hip joint tuberculosis: complications during treatment of disseminated isoniazid-resistant tuberculosis in an immunocompromised host. Journal of Clinical Microbiology. 2009;47(2):507–10.

23. Pranger AD, Alffenaar J-WC, Wessels AMA, Greijdanus B, Uges DRA. Determination of moxifloxacin in human plasma, plasma ultrafiltrate, and cerebrospinal fluid by a rapid and simple liquid chromatography-tandem mass spectrometry method. J Anal Toxicol. 2010;34(3):135–41.

24. Alffenaar JWC, de Vries PM, Luijckx GJ, van Soolingen D, van der Werf TS, van Altena R. Plasma and cerebrospinal fluid pharmacokinetics of moxifloxacin in a patient with tuberculous meningitis. Antimicrob Agents Chemother. 2008;52(6):2293–5.

25. Uchida Y, Zhang Z, Tachikawa M, Terasaki T. Quantitative targeted absolute proteomics of rat blood-cerebrospinal fluid barrier transporters: comparison with a human specimen. J Neurochem. 2015;134(6):1104–15.

26. Kubo Y, Ohtsuki S, Uchida Y, Terasaki T. Quantitative Determination of Luminal and Abluminal Membrane Distributions of Transporters in Porcine Brain Capillaries by Plasma Membrane Fractionation and Quantitative Targeted Proteomics. J Pharm Sci. 2015;104(9):3060–8.

27. Uchida Y, Goto R, Takeuchi H, Łuczak M, Usui T, Tachikawa M, et al. Abundant Expression of OCT2, MATE1, OAT1, OAT3, PEPT2, BCRP, MDR1, and xCT Transporters in Blood-Arachnoid Barrier of Pig and Polarized Localizations at CSF- and Blood-Facing Plasma Membranes. Drug Metab Dispos. 2020;48(2):135–45.

28. Seymour RS, Angove SE, Snelling EP, Cassey P. Scaling of cerebral blood perfusion in primates and marsupials. J Exp Biol. 2015;218(Pt 16):2631–40.

29. Wong AD, Ye M, Levy AF, Rothstein JD, Bergles DE, Searson PC. The blood-brain barrier: an engineering perspective. Front Neuroeng. 2013;6:7.

30. Zhanel GG, Ennis K, Vercaigne L, Walkty A, Gin AS, Embil J, et al. A critical review of the fluoroquinolones: focus on respiratory infections. Drugs. 2002;62(1):13–59.

31. Siefert HM, Domdey-Bette A, Henninger K, Hucke F, Kohlsdorfer C, Stass HH. Pharmacokinetics of the 8-methoxyquinolone, moxifloxacin: a comparison in humans and other mammalian species. J Antimicrob Chemother. 1999;43 Suppl B:69-76.

32. Vogensen VB, Bolhuis MS, Sturkenboom MGG, van der Werf TS, de Lange WCM, Anthony RM, et al. Clinical Relevance of Rifampicin-Moxifloxacin Interaction in Isoniazid-Resistant/Intolerant Tuberculosis Patients. Antimicrob Agents Chemother. 2022;66(2):e0182921.

33. van den Elsen SHJ, Sturkenboom MGG, Akkerman OW, Manika K, Kioumis IP, van der Werf TS, et al. Limited Sampling Strategies Using Linear Regression and the Bayesian Approach for Therapeutic Drug Monitoring of Moxifloxacin in Tuberculosis Patients. Antimicrob Agents Chemother. 2019;63(7).

34. Nguyen THT, Mouksassi M-S, Holford N, Al-Huniti N, Freedman I, Hooker AC, et al. Model Evaluation of Continuous Data Pharmacometric Models: Metrics and Graphics. CPT: Pharmacometrics & Systems Pharmacology. 2017;6(2):87–109.

35. Nuermberger E, Grosset J. Pharmacokinetic and pharmacodynamic issues in the treatment of mycobacterial infections. Eur J Clin Microbiol Infect Dis. 2004;23(4):243–55.

36. Gumbo T, Louie A, Deziel MR, Parsons LM, Salfinger M, Drusano GL. Selection of a moxifloxacin dose that suppresses drug resistance in Mycobacterium tuberculosis, by use of an in vitro pharmacodynamic infection model and mathematical modeling. J Infect Dis. 2004;190(9):1642–51.

37. Angeby KA, Jureen P, Giske CG, Chryssanthou E, Sturegård E, Nordvall M, et al. Wild-type MIC distributions of four fluoroquinolones active against Mycobacterium tuberculosis in relation to current critical concentrations and available pharmacokinetic and pharmacodynamic data. J Antimicrob Chemother. 2010;65(5):946–52.

38. Simulations Plus. Monolix. 2024.

39. Wang W, Hallow KM, James DA. A Tutorial on RxODE: Simulating Differential Equation Pharmacometric Models in R. CPT Pharmacometrics Syst Pharmacol. 2016;5(1):3–10.

40. Hosmann A, Moser MM, van Os W, Gramms L, Al Jalali V, Sanz Codina M, et al. Linezolid brain penetration in neurointensive care patients. J Antimicrob Chemother. 2024;79(3):669–77.

41. Chauzy A, Bouchène S, Aranzana-Climent V, Clarhaut J, Adier C, Grégoire N, et al. A Minimal Physiologically Based Pharmacokinetic Model to Characterize CNS Distribution of Metronidazole in Neuro Care ICU Patients. Antibiotics. 2022;11(10):1293.

42. Westerhout J, van den Berg DJ, Hartman R, Danhof M, de Lange EC. Prediction of methotrexate CNS distribution in different species - influence of disease conditions. Eur J Pharm Sci. 2014;57:11–24.

43. Shapiro WR, Young DF, Mehta BM. Methotrexate: distribution in cerebrospinal fluid after intravenous, ventricular and lumbar injections. N Engl J Med. 1975;293(4):161–6.

44. Pioget JC, Wolff M, Singlas E, Laisne MJ, Clair B, Regnier B, et al. Diffusion of ofloxacin into cerebrospinal fluid of patients with purulent meningitis or ventriculitis. Antimicrob Agents Chemother. 1989;33(6):933–6.

45. Nau R, Kinzig M, Dreyhaupt T, Kolenda H, Sörgel F, Prange HW. Kinetics of ofloxacin and its metabolites in cerebrospinal fluid after a single intravenous infusion of 400 milligrams of ofloxacin. Antimicrob Agents Chemother. 1994;38(8):1849–53.

46. Ruslami R, Ganiem AR, Dian S, Apriani L, Achmad TH, van der Ven AJ, et al. Intensified regimen containing rifampicin and moxifloxacin for tuberculous meningitis: an open-label, randomised controlled phase 2 trial. Lancet Infect Dis. 2013;13(1):27–35.

47. van Haarst A, Smith S, Garvin C, Benrimoh N, Paglialunga S. Rifampin Drug-Drug-Interaction Studies: Reflections on the Nitrosamine Impurities Issue. Clin Pharmacol Ther. 2023;113(4):816–21.

48. Keating GM, Scott □. Moxifloxacin: a review of its use in the management of bacterial infections. Drugs. 2004;64(20):2347–77.

49. Kuban W, Daniel WA. Cytochrome P450 expression and regulation in the brain. Drug Metabolism Reviews. 2021;53(1).

50. Decleves X, Jacob A, Yousif S, Shawahna R, Potin S, Scherrmann J-M. Interplay of drug metabolizing CYP450 enzymes and ABC transporters in the blood-brain barrier. Curr Drug Metab. 2011;12(8):732–41.

51. Inbaraj LR, Manesh A, Ponnuraja C, Bhaskar A, Srinivasalu VA, Daniel BD. Comparative evaluation of intensified short course regimen and standard regimen for adults TB meningitis: a protocol for an open label, multi-center, parallel arms, randomized controlled superiority trial (INSHORT trial). Trials. 2024;25(1):294.

52. Manyelo CM, Solomons RS, Walzl G, Chegou NN. Tuberculous Meningitis: Pathogenesis, Immune Responses, Diagnostic Challenges, and the Potential of Biomarker-Based Approaches. J Clin Microbiol. 2021;59(3).

53. Ball K, Bouzom F, Scherrmann JM, Walther B, Declèves X. Physiologically based pharmacokinetic modelling of drug penetration across the blood-brain barrier--towards a mechanistic IVIVE-based approach. Aaps j. 2013;15(4):913–32.

54. Yamamoto Y, Danhof M, de Lange ECM. Microdialysis: the Key to Physiologically Based Model Prediction of Human CNS Target Site Concentrations. The AAPS Journal. 2017;19(4):891–909.

55. Cserr HF. Physiology of the choroid plexus. Physiol Rev. 1971;51(2):273–311.

56. Bessen MA, Gayen CD, Quarrington RD, Walls AC, Leonard AV, Kurtcuoglu V, et al. Characterising spinal cerebrospinal fluid flow in the pig with phase-contrast magnetic resonance imaging. Fluids and Barriers of the CNS. 2023;20(1):5.

57. Chun S-W, Lee H-J, Nam K-H, Sohn C-H, Kim KD, Jeong E-J, et al. Cerebrospinal fluid dynamics at the lumbosacral level in patients with spinal stenosis: A pilot study. J Orthop Res. 2017;35(1):104–12.

58. Hirasawa M, de Lange ECM. Revisiting Cerebrospinal Fluid Flow Direction and Rate in Physiologically Based Pharmacokinetic Model. Pharmaceutics. 2022;14(9).

59. Hladky SB, Barrand MA. Mechanisms of fluid movement into, through and out of the brain: evaluation of the evidence. Fluids Barriers CNS. 2014;11(1):26.

60. Klarica M, Radoš M, Orešković D. The Movement of Cerebrospinal Fluid and Its Relationship with Substances Behavior in Cerebrospinal and Interstitial Fluid. Neuroscience. 2019;414:28–48.

61. MacAulay N, Keep RF, Zeuthen T. Cerebrospinal fluid production by the choroid plexus: a century of barrier research revisited. Fluids and Barriers of the CNS. 2022;19(1):26.

62. van Valkengoed DW, Krekels EHJ, Knibbe CAJ. All You Need to Know About Allometric Scaling: An Integrative Review on the Theoretical Basis, Empirical Evidence, and Application in Human Pharmacology. Clinical Pharmacokinetics. 2024.

63. Barber TW, Brockway JA, Higgins LS. The density of tissues in and about the head. Acta Neurol Scand. 1970;46(1):85–92.

64. DiResta GR, Lee J, Lau N, Ali F, Galicich JH, Arbit E, editors. Measurement of Brain Tissue Density Using Pycnometry. Brain Edema VIII; 1990 1990//; Vienna: Springer Vienna.

65. Boutaleb S, Pouget JP, Hindorf C, Pelegrin A, Barbet J, Kotzki PO, et al. Impact of Mouse Model on Preclinical Dosimetry in Targeted Radionuclide Therapy. Proceedings of the IEEE. 2009;97(12):2076–85.

66. Karbowski J. Global and regional brain metabolic scaling and its functional consequences. BMC Biol. 2007;5:18.

67. Stass H, Dalhoff A, Kubitza D, Schühly U. Pharmacokinetics, safety, and tolerability of ascending single doses of moxifloxacin, a new 8-methoxy quinolone, administered to healthy subjects. Antimicrob Agents Chemother. 1998;42(8):2060–5.

68. Paliwal VK, Garg RK. Hydrocephalus in Tuberculous Meningitis - Pearls and Nuances. Neurology India. 2021;69(Supplement):S330–S5.

69. Drusano GL, Sgambati N, Eichas A, Brown DL, Kulawy R, Louie A. The combination of rifampin plus moxifloxacin is synergistic for suppression of resistance but antagonistic for cell kill of Mycobacterium tuberculosis as determined in a hollow-fiber infection model. mBio. 2010;1(3).

70. Conrad MS, Dilger RN, Johnson RW. Brain growth of the domestic pig (Sus scrofa) from 2 to 24 weeks of age: a longitudinal MRI study. Developmental Neuroscience. 2012;34(4):291–8.

71. Fil JE, Joung S, Zimmerman BJ, Sutton BP, Dilger RN. High-resolution magnetic resonance imaging-based atlases for the young and adolescent domesticated pig (Sus scrofa). Journal of Neuroscience Methods. 2021;354:109107.

72. George I, Fawehinmi H, Oyakhire M, Musa S, Akintola O. COMPARATIVE STUDIES ON THE BRAINS OF LOCAL BREEDS OF PIG (LANDRACE BREED) AND DOG (MONGREL BREED). European Journal of Biomedical. 2020;7(1):324–30.

73. Rushton JP, Ankney CD. Whole brain size and general mental ability: a review. Int J Neurosci. 2009;119(5):691–731.

74. Ungerstedt U. Microdialysis--principles and applications for studies in animals and man. J Intern Med. 1991;230(4):365–73.

75. Lei Y, Han H, Yuan F, Javeed A, Zhao Y. The brain interstitial system: Anatomy, modeling, in vivo measurement, and applications. Progress In Neurobiology. 2017;157:230–46.

76. Morgan CJ, Pyne-Geithman GJ, Jauch EC, Shukla R, Wagner KR, Clark JF, et al. Bilirubin as a cerebrospinal fluid marker of sentinel subarachnoid hemorrhage: a preliminary report in pigs. J Neurosurg. 2004;101(6):1026–9.

77. Proulx ST. Cerebrospinal fluid outflow: a review of the historical and contemporary evidence for arachnoid villi, perineural routes, and dural lymphatics. Cell Mol Life Sci. 2021;78(6):2429–57.

78. Mayfrank L, Kissler J, Raoofi R, Delsing P, Weis J, Küker W, et al. Ventricular dilatation in experimental intraventricular hemorrhage in pigs. Characterization of cerebrospinal fluid dynamics and the effects of fibrinolytic treatment. Stroke. 1997;28(1):141–8.

79. Jacob RM, Mudd AT, Alexander LS, Lai CS, Dilger RN. Comparison of Brain Development in Sow-Reared and Artificially Reared Piglets. Front Pediatr. 2016;4:95.

80. Vikner T, Johnson KM, Cadman RV, Betthauser TJ, Wilson RE, Chin N, et al. CSF dynamics throughout the ventricular system using 4D flow MRI: associations to arterial pulsatility, ventricular volumes, and age. Fluids Barriers CNS. 2024;21(1):68.

81. Romagnoli N, Ventrella D, Giunti M, Dondi F, Sorrentino NC, Fraldi A, et al. Access to cerebrospinal fluid in piglets via the cisterna magna: optimization and description of the technique. Lab Anim. 2014;48(4):345–8.

82. Jakola AS, Jørgensen A, Selbekk T, Michler RP, Solheim O, Torp SH, et al. Animal study assessing safety of an acoustic coupling fluid that holds the potential to avoid surgically induced artifacts in 3D ultrasound guided operations. BMC Med Imaging. 2014;14:11.

83. Rios JC, Galper MW, Naidich TP. CHAPTER 13 - Ventricles and Intracranial Subarachnoid Spaces. In: Naidich TP, Castillo M, Cha S, Smirniotopoulos JG, editors. Imaging of the Brain. Philadelphia: W.B. Saunders; 2013. p. 245–71.

84. Dicke U, Roth G. Neuronal factors determining high intelligence. Philos Trans R Soc Lond B Biol Sci. 2016;371(1685):20150180.

85. Spector R, Keep RF, Robert Snodgrass S, Smith QR, Johanson CE. A balanced view of choroid plexus structure and function: Focus on adult humans. Exp Neurol. 2015;267:78–86.

86. Soltan N, Siegmund GP, Cripton PA, Jones CF. Geometric and Inertial Properties of the Pig Head and Brain in an Anatomical Coordinate System. Ann Biomed Eng. 2023;51(11):2544–53.

87. Anwar M, Weiss J, Weiss HR. Quantitative determination of morphometric indices of the total and perfused capillary network of the newborn pig brain. Pediatr Res. 1992;32(5):542–6.

88. Kawai R, Mathew D, Tanaka C, Rowland M. Physiologically based pharmacokinetics of cyclosporine A: extension to tissue distribution kinetics in rats and scale-up to human. The Journal of Pharmacology and Experimental Therapeutics. 1998;287(2):457–68.

